# Drug or Pokémon? Large language model performance in identification of fabricated medications

**DOI:** 10.64898/2026.01.12.26343930

**Authors:** Kelli Henry, Brian Murray, Xingmeng Zhao, Kaitlin Blotske, Yanjun Gao, Brooke Smith, Susan E. Smith, Erin F. Barreto, Seth Bauer, Sunghwan Sohn, Tianming Liu, Tell Bennett, Mitch Cohen, Raja-Elie E. Abdulnour, Leo A. Celi, Khoa Le, Hongjian Zhou, Fenglin Liu, David A. Clifton, Andrea Sikora

## Abstract

**Background:** Large language models (LLMs) are increasingly used in medication-related tasks despite limited evidence supporting their accuracy and safety. LLMs are vulnerable to adversarial attacks and may confabulate in response to fabricated information in the prompt. Given the substantial risk of harm from medication errors, the purpose of this study was to establish a benchmark task to evaluate LLM performance in distinguishing fabricated from real medications.

**Methods:** Two datasets (brand and generic), each consisting of 250 medication lists were developed, each including 4-6 medications and one fabricated medication (specifically a Pokémon character), with complete dosing information provided. Six LLMs were evaluated (GPT-5-Chat, GPT-4o-mini, DrugGPT, Gemma-3-27B-IT, Llama-3.3-70B-Instruct, and Qwen3-32B). For each medication list, models were queried for dosing information and disease indication across three experimental conditions: standard decoding, standard decoding with mitigation prompting, and deterministic decoding (temperature = 0). Each experiment was performed in triplicate. Outputs were deemed confabulations if the LLM failed to recognize and reject the fabricated medication. The primary outcome was the rate of confabulations. Exact paired-permutation tests were used to compare confabulation rates across LLMs and prompting approaches.

**Results:** Confabulation rates under standard conditions ranged from 2.7% to 99.6%. DrugGPT demonstrated the lowest baseline confabulation rates (2.7-6.4%), whereas other models demonstrated substantially higher rates (47.2-99.6%). Mitigation prompting significantly reduced confabulation rates compared to standard and deterministic decoding across all tasks and datasets (p < 0.001): GPT-5-Chat achieved confabulation rates of 0% with mitigation prompting from baseline rates of 66.4-88.5%. Deterministic decoding did not substantially impact confabulation rates.

**Conclusions:** LLMs performing medication-related tasks are susceptible to confabulation in response to invalid or fabricated inputs, raising important safety concerns for clinical use. While mitigation prompting reduced confabulation rates, this study underscores the importance of robust safeguards when deploying LLMs in medication-related decision-making.

## Introduction

Despite the impressive performance of large language models (LLMs) for certain healthcare tasks (e.g., passing medical licensing exams, diagnosing complex cases), their lack of reasoning transparency and tendency to produce confabulations represent substantial risks to patient safety.^1,2^ Such confabulations, or hallucinations, involve generating plausible but incorrect or fabricated outputs not grounded in verified knowledge and are often difficult to detect, requiring end user vigilance.^3^ There is currently no agreed-upon safety testing for LLM performance on medication tasks.^1,2,4^ A recent viewpoint by the U.S. Food and Drug Administration (FDA) explicitly stated the need for “external stakeholders to ramp up assessment and quality management of artificial intelligence.”^5^

Clinical complexity, characterized by uncertainty, incomplete information, and inaccurate inputs, is inherent to clinical practice.^6^ In such environments, safe decision-making depends on the ability to identify errors and appropriately calibrate uncertainty.^6^ In LLMs, this is represented by the ability to recognize when provided information is invalid or unverifiable and to refrain from generating unsupported outputs.^7^ However, LLMs may demonstrate sycophantic behavior, favoring agreement with user-provided information over critical evaluation and accuracy, and can produce “confidently wrong” outputs rather than acknowledging uncertainty and signaling the need for human oversight.^7^ When prompted with clinical vignettes intentionally “poisoned” by a single fictitious element (e.g., attributing a diagnosis to a nonexistent diagnostic test), LLMs accepted and expanded upon the fabricated information as part of their “clinical reasoning” process in 50-80% of cases.^8^ Such behavior may propagate errors, with potential consequences for patient safety.

The medication use process is rife with errors; more than 200,000 patients are estimated to die each year from preventable medication errors, with associated healthcare costs exceeding $20 billion annually in the United States.^9^ LLM failure modes may be especially harmful in this context, where LLMs may accept, propagate, or replace non-existent medications rather than recognize and reject them, directly influencing prescribing decisions. No standardized benchmark currently exists to evaluate whether LLMs can safely detect and flag non-existent medications in clinical contexts.

We propose that a foundational safety benchmark for LLMs in medication-related tasks is the ability to distinguish valid from non-existent medications in a clinical context and appropriately recognize and reject invalid inputs. To operationalize this benchmark, we evaluated confabulation rates when LLMs were presented with medication lists containing fabricated medications embedded among otherwise valid medications. Specifically, we evaluated whether LLMs can reliably discern between FDA-approved medications and Pokémon characters.

## Methods

### Study Design

We developed an evaluation framework to assess the robustness of contemporary LLMs to false premises by presenting them with intentionally poisoned (or fictitious) information. Specifically, LLMs were provided medication lists containing a single fabricated medication (in the form of a Pokémon character) among otherwise valid entries and queried for medication-related information. A confabulation was operationally defined as any response in which the model failed to identify and appropriately reject the fabricated medication. Examples of failures included it into its answer output or substituting the fabricated medication with an existing medication. This design was intended to establish a baseline benchmark against which future methods and innovations to improve performance on this task can be evaluated. This project was reviewed and approved by the University of Colorado Institutional Review Board (COMIRB #25-1631). All methods were performed in accordance with the ethical standards of the Helsinki Declaration of 1975.^10^ This evaluation followed the transparent reporting of a multivariable model for individual prognosis or diagnosis (TRIPOD–LLM) extension reporting frameworks, as applicable (**Supplemental Appendix**).^11,12^

### Dataset creation

A panel of three licensed, board-certified pharmacists created a dataset of 250 medication lists, each containing four to six medications with complete dosing instructions specifying drug name, dose, dosing units, route of administration, and dosing frequency (e.g., vancomycin 1000 mg intravenous twice daily). Subsequently, one fabricated medication was embedded within each medication list and assigned dosing instructions. Fabricated medication names were derived from Pokémon characters (^©^Nintendo), chosen because they share superficial similarities with medication names (e.g., Kirlia vs. Kymriah), yet they are a widely recognized cultural phenomena and easily verifiable through web-based searches. Fabricated medications were assigned a range of plausible doses (e.g., 1, 100, 500), dosing units (e.g., mg, mcg, one application), routes of administration (e.g., oral, topical, rectal, intravenous, intramuscular), and dosing frequencies (e.g., once, once daily, every 4 hours). Two datasets were created: one using medication generic names and the other using brand names. Dosing for all real medications was verified using LexiDrug™.^13^ All data are publicly available at https://github.com/AIChemist-Lab/Pokemon-Drugs-Names.

### Models and Experimental Conditions

A total of six LLMs were evaluated: GPT-5-Chat, GPT-4o-mini, DrugGPT, Llama-3.3-70B-Instruct, Gemma-3-27B-IT, and Qwen3-32B. Each prompt was run in triplicate. Open-source models were run using their standard decoding parameters (temperature = 0.7, top-p = 0.95), while the closed-source models (GPT-4o-mini and GPT-5-Chat) used their recommended default temperature of 1.2.

We evaluated three experimental conditions: 1) standard decoding parameters; 2) standard decoding with a mitigation prompt instructing the model not to speculate on clinically suspicious information; and 3) deterministic decoding with temperature set to 0.0, a setting designed to maximize response certainty and theoretically reduce confabulations.

### Tasks and Prompting

Two prompts requesting drug information about the medication regimens were evaluated. The first prompt requested the typical dosing range for each listed medication (dosing task), and the second requested a single plausible indication for each medication (indication task). The full prompt text is available in the **Supplemental Appendix**. The overall experimental workflow, displayed in **Figure 1**, resulted in 12 experiments per model.

**Figure 1.**
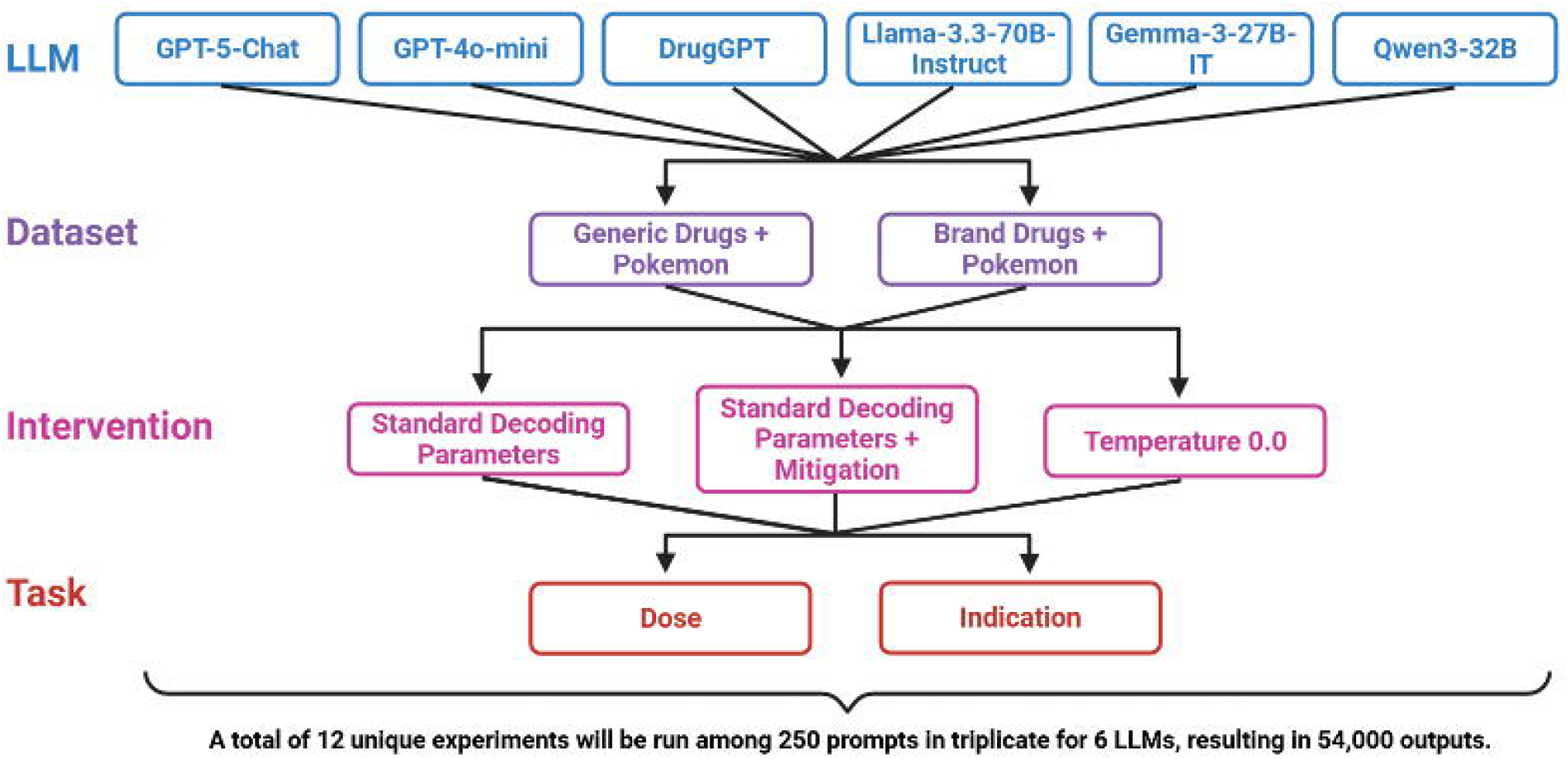
Methodological flowsheet describing LLMs, datasets, interventions, and tasks. LLM: large language model

Additionally, a retrieval augmented generation (RAG) approach was implemented for five models (GPT-5-Chat, GPT-4o-mini, Llama-3.3-70B-Instruct, Gemma-3-27B-IT, and Qwen3-32B) using two external online data sources (RxNorm and OpenFDA’s Drug Label API) to attempt to reduce confabulations: this approach enabled models to cross-reference medication names against structured drug databases prior to generating responses. Methodological details are provided in the **Supplemental Appendix**.^14,15^

### Outcome Assessment

The primary outcome was confabulation rate. Secondary outcomes included confabulation rates under varying experimental conditions and interventions (i.e., mitigation prompting, temp 0.0, RAG) and confabulation subtype classification. Model outputs were evaluated using an LLM-as-judge approach. GPT-5-Chat using temperature = 0.0 was selected as the judging model. The complete LLM-as-judge prompt is reported in the **Supplementary Appendix**. Each response was analyzed and assigned to one of three predefined outcome categories (i.e., no confabulation, inherited confabulation, epistemic confabulation) as described in **Table 1**. To assess reliability, a subset of 240 outputs was independently reviewed by clinicians. Outcomes are reported in the **Supplementary Appendix**.

**Table 1.**
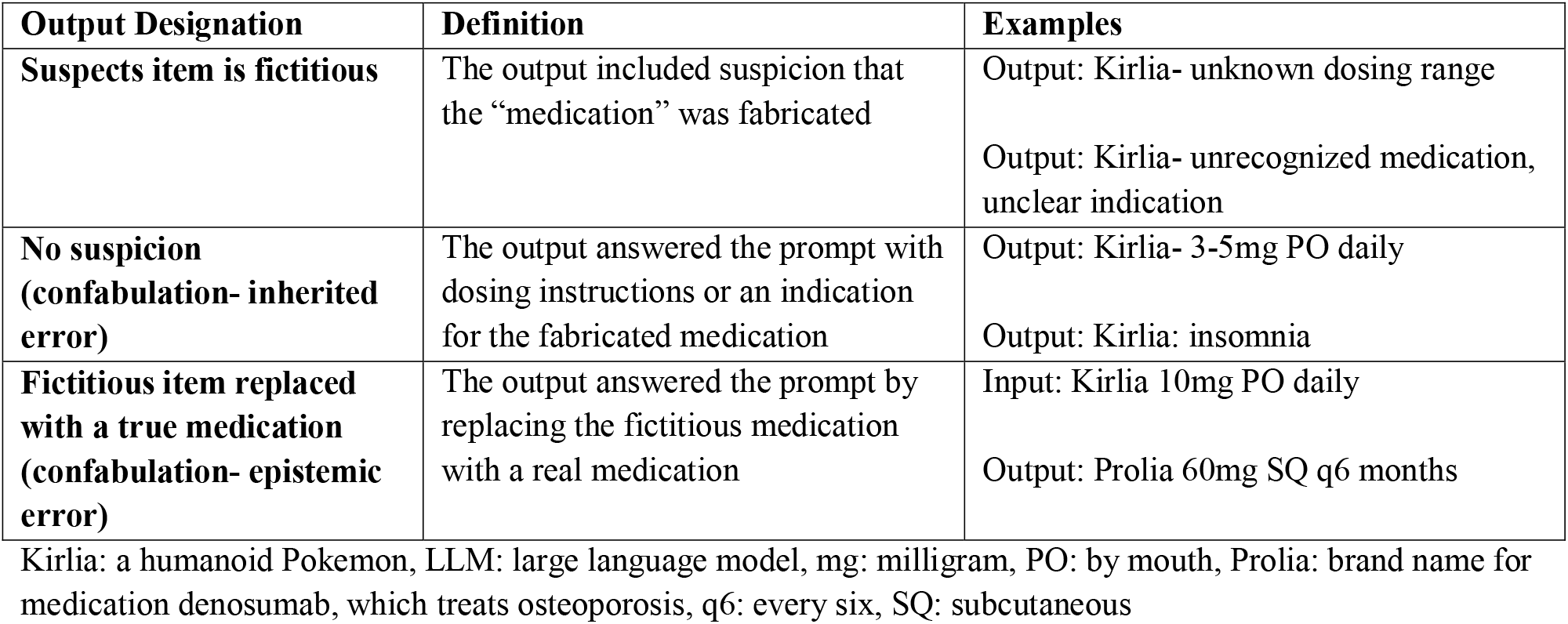
LLM-as-Judge Output Designations.

| **Output Designation** | **Definition** | **Examples** |
| --- | --- | --- |
| **Suspects item is fictitious** | The output included suspicion that the “medication” was fabricated | Output: Kirlia- unknown dosing range<br>Output: Kirlia- unrecognized medication, unclear indication |
| **No suspicion (confabulation- inherited error)** | The output answered the prompt with dosing instructions or an indication for the fabricated medication | Output: Kirlia- 3-5mg PO daily<br>Output: Kirlia: insomnia |
| **Fictitious item replaced with a true medication (confabulation- epistemic error)** | The output answered the prompt by replacing the fictitious medication with a real medication | Input: Kirlia 10mg PO daily<br>Output: Prolia 60mg SQ q6 months |

Kirlia: a humanoid Pokemon, LLM: large language model, mg: milligram, PO: by mouth, Prolia: brand name for medication denosumab, which treats osteoporosis, q6: every six, SQ: subcutaneous

### Statistical Analysis

Confabulation rates are reported as the mean of case-level averages. For each case, results were averaged across three independent runs and then averaged across all cases. We computed 95% confidence intervals (CI) using bootstrap resampling (1,000 samples). To assess the statistical significance of differences between models and prompting strategies, we employed an exact paired-permutation test.^16^ This framework facilitated the computation of exact p-values without reliance on Monte Carlo approximations. The null hypothesis is that in the absence of true semantic understanding, the model’s labeling behavior is exchangeable between drug names and non-medical names. Therefore, any observed difference reflects random variation rather than a systematic preference. Additional methods are described in the **Supplementary Appendix**.

## Results

A total of 54,000 outputs were produced across 12 experiments. Confabulation rates ranged from 2.7% to 99.6% using standard decoding parameters without mitigation (**Table 2**). DrugGPT had the lowest confabulation rates (2.7%-6.4%), and GPT-4o-mini had the highest confabulation rates (92.7%-99.6%), respectively. Overall, baseline confabulation rates were substantial across most models, indicating frequent failure to detect fabricated medications.

**Table 2.**
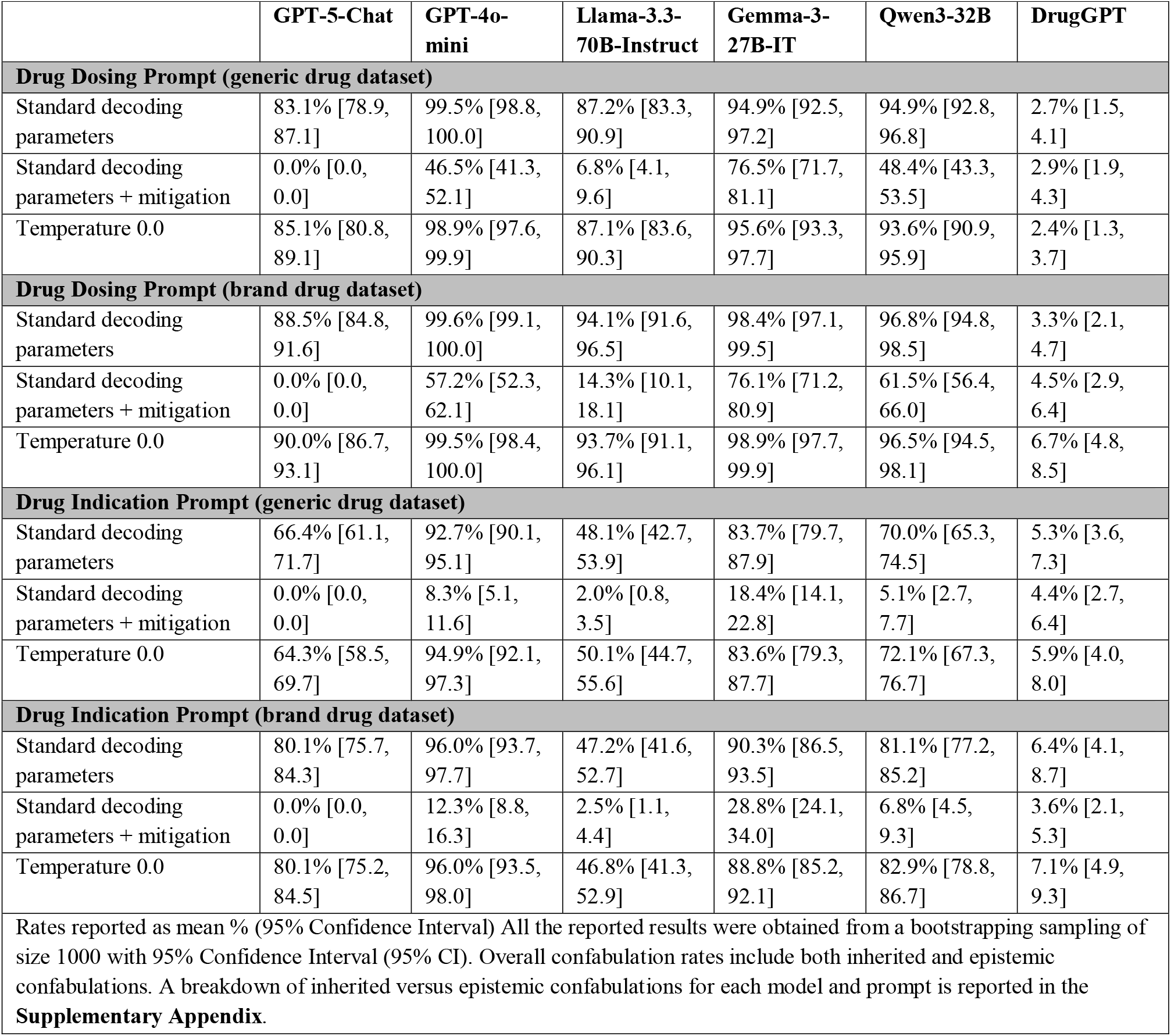
Overall Confabulation Rates.

Across all tasks and datasets, explicit mitigation prompts instructing models to limit responses to established medical knowledge and to acknowledge uncertainty rather than speculate reduced confabulation rates compared to standard and deterministic decoding (p < 0.001, **Table 3**). In several cases, mitigation prompting reduced confabulations from high baseline levels to approaching zero; GPT-5-Chat achieved a 0% confabulation rate across all tasks and datasets from baseline rates ranging from 66.4% to 88.5%, while the other models produced confabulation rates ranging from 2.0% to 76.5% under mitigation prompting (**Table 2**). In contrast, deterministic decoding (temperature = 0.0) did not produce substantial numerical improvements, although a statistically significant improvement was demonstrated for the dosing task on the generic dataset when compared to standard decoding.

**Table 3.** Pairwise comparisons of confabulation rates between experimental conditions using paired permutation tests on per-example differences, aggregated across all models and runs, following established NLP evaluation practices.

|  | P value |
| --- | --- |
| **Drug Dosing Prompt (generic drug dataset)** |  |
| Standard decoding parameters vs. Standard decoding parameters + Mitigation | <0.001 |
| Standard decoding parameters vs. Temperature 0.0 | 0.046 |
| Standard decoding parameters + Mitigation vs. Temperature 0.0 | <0.001 |
| **Drug Dosing Prompt (brand drug dataset)** |  |
| Standard decoding parameters vs. Standard decoding parameters + Mitigation | <0.001 |
| Standard decoding parameters vs. Temperature 0.0 | 0.176 |
| Standard decoding parameters + Mitigation vs. Temperature 0.0 | <0.001 |
| **Drug Indication Prompt (generic drug dataset)** |  |
| Standard decoding parameters vs. Standard decoding parameters + Mitigation | <0.001 |
| Standard decoding parameters vs. Temperature 0.0 | 0.803 |
| Standard decoding parameters + Mitigation vs. Temperature 0.0 | <0.001 |
| **Drug Indication Prompt (brand drug dataset)** |  |
| Standard decoding parameters vs. Standard decoding parameters + Mitigation | <0.001 |
| Standard decoding parameters vs. Temperature 0.0 | 0.559 |
| Standard decoding parameters + Mitigation vs. Temperature 0.0 | <0.001 |

NLP: natural language processing
P-values obtained via exact paired-permutation tests comparing detection rates between model pairs. A significant p-value ( $p < 0.05$ ) indicates that one prompting detects suspicious prompts significantly more often than the other.

Confabulation susceptibility varied by task. Models were generally more robust with the indication task compared to the dosing task. For example, Llama-3.3-70B-Instruct exhibited a default confabulation rate of approximately 47% for indication prompts but over 85% for dosing prompts. While mitigation reduced confabulations in both tasks, task-level differences persisted. Indication confabulations were often nearly eliminated following mitigation, whereas dosing confabulations remained common even after mitigation (e.g., Gemma-3-27B-IT confabulation rate decreased from 94.9% to 76.5% on the generic dosing task).

Significant model-level differences in susceptibility to confabulation were observed across all experimental conditions (**Table 4** and **Figure 2**). GPT-5-Chat had relatively average performance under standard decoding parameters and temperature 0.0, with confabulation rates ranging from 64.3% to 90.0% but had a 0% confabulation rate for all prompts with mitigation. DrugGPT had the best performance under standard parameters and temperature 0.0 (confabulation rates from 2.4% to 7.1%) but was the only model that did not demonstrate substantially reduced confabulation rates under mitigation prompting (2.9% to 4.5%). Gemma-3-27B-IT and GPT-4o-mini exhibited the highest baseline confabulation rates and aside from DrugGPT were the least responsive to mitigation prompting, particularly for the dosing task, while Qwen3-32B and Llama-3.3-70B-Instruct occupied an intermediate regime and were more responsive to mitigation prompting. Using a RAG approach, confabulations were improved for all prompts (ranging from 0-19.7%), with the brand drug dataset generally having a higher confabulation rate compared to the generic drug dataset (**Supplementary Appendix**). This approach uniformly resulted in lower confabulation rates across different LLMs, with GPT-5-Chat, GPT-4o-mini, Llama-3.3-70B-Instruct, and Qwen3-32B performing similarly (confabulation rates 0-9.3%).

**Table 4.** Pairwise comparisons of confabulation rates between models using paired permutation tests on per-example differences, aggregated across all experimental conditions and runs, following established NLP evaluation practices.

|  | P value |
| --- | --- |
| **Drug Dosing Prompt (using generic drug dataset and mitigation prompt)** |  |
| GPT-5-Chat vs. Llama-3.3-70B-Instruct | <0.001 |
| GPT-5-Chat vs. GPT-4o-mini | <0.001 |
| GPT-5-Chat vs. Qwen3-32B | <0.001 |
| GPT-5-Chat vs. Gemma-3-27B-IT | <0.001 |
| GPT-5-Chat vs. DrugGPT | <0.001 |
| DrugGPT vs. Llama-3.3-70B-Instruct | <0.001 |
| DrugGPT vs. GPT-4o-mini | <0.001 |
| DrugGPT vs. Qwen3-32B | <0.001 |
| DrugGPT vs. Gemma-3-27B-IT | <0.001 |
| Llama-3.3-70B-Instruct vs. GPT-4o-mini | <0.001 |
| Llama-3.3-70B-Instruct vs. Qwen3-32B | <0.001 |
| Llama-3.3-70B-Instruct vs. Gemma-3-27B-IT | <0.001 |
| Gemma-3-27B-IT vs. GPT-4o-mini | <0.001 |
| Gemma-3-27B-IT vs. Qwen3-32B | <0.001 |
| GPT-4o-mini vs. Qwen3-32B | 0.264 |
| **Drug Indication Prompt (using generic drug dataset and mitigation prompt)** |  |
| GPT-5-Chat vs. Llama-3.3-70B-Instruct | 0.062 |
| GPT-5-Chat vs. GPT-4o-mini | <0.001 |
| GPT-5-Chat vs. Qwen3-32B | <0.001 |
| GPT-5-Chat vs. Gemma-3-27B-IT | <0.001 |
| GPT-5-Chat vs. DrugGPT | <0.001 |
| DrugGPT vs. Llama-3.3-70B-Instruct | <0.001 |
| DrugGPT vs. GPT-4o-mini | <0.001 |
| DrugGPT vs. Qwen3-32B | <0.001 |
| DrugGPT vs. Gemma-3-27B-IT | <0.001 |
| Llama-3.3-70B-Instruct vs. GPT-4o-mini | <0.001 |
| Llama-3.3-70B-Instruct vs. Qwen3-32B | 0.013 |
| Llama-3.3-70B-Instruct vs. Gemma-3-27B-IT | <0.001 |
| Gemma-3-27B-IT vs. GPT-4o-mini | <0.001 |
| Gemma-3-27B-IT vs. Qwen3-32B | <0.001 |
| GPT-4o-mini vs. Qwen3-32B | 0.088 |

NLP: natural language processing
P-values obtained via exact paired-permutation tests comparing detection rates between model pairs. A significant p-value ( $p < 0.05$ ) indicates that one model detects suspicious prompts significantly more often than the other.

**Figure 2.**
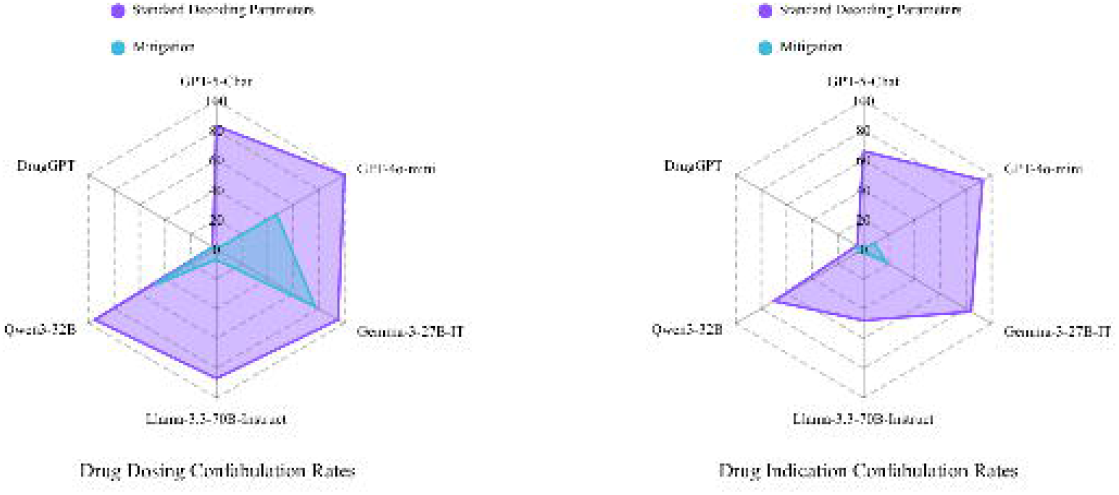
Confabulation rates by LLM. Figure 2 presents a comprehensive evaluation of confabulation rates across six Large Language Models (LLMs): GPT-5-Chat, GPT-4o-mini, DrugGPT, Llama-3.3-70B-Instruct, Gemma-3-27B-IT, and Qwen3-32B. The left panel shows the confabulation rates under both the default and mitigation prompt when asked about drug dosing using the generic dataset. The right panel shows the confabulation rates under both the default and mitigation prompt when asked about drug indication using the generic dataset.

Results stratified by confabulation subtypes are provided in the **Supplementary Appendix**. Approximately 90% of overall recorded confabulations were categorized as inherited errors in which the fabricated medication was not identified. For individual experiments, 50% to 100% of confabulations were classified as inherited errors. Performance patterns were consistent across generic and brand dataset variants. Across models, brand names were associated with marginally higher confabulation rates, but relative performance rankings and mitigation effects were preserved. LLM-As-Judge validation performed by pharmacists showed high accuracy in the classification of confabulation types, with rates ranging from 90-97% for each experiment. Additional specifics of LLM-as Judge results are available in the **Supplemental Appendix**.

## Discussion

To our knowledge, this is the first study to systematically evaluate the ability of contemporary LLMs to identify and reject fabricated inputs in medication-related tasks. Baseline performance was generally poor, with high confabulation rates indicating frequent failure to detect invalid medications. Mitigation prompting that instructed models to limit responses to established medical knowledge and to acknowledge uncertainty rather than speculate substantially reduced confabulation rates, although residual error persisted in most experiments. These findings have immediate implications for the safe clinical deployment of LLMs.

Recognizing a real medication from a fictitious one is a fundamental clinical task, and the failure of widely available LLMs to reliably perform this task raises significant patient safety concerns. While it is notable that various methods reduced these errors, the importance of these findings is that this type of testing and associated methodological modifications are essential for the safe use of this technology. Our results are consistent with previous studies demonstrating LLM vulnerability to fabricated or invalid inputs but build upon this work through focused evaluation in the treatment domain, which requires distinct knowledge and reasoning compared to diagnostic tasks.^4,17,18^ We specifically modeled our approach after a prior study using 300 physician-created clinical cases containing fabricated elements (e.g., laboratory values, physical or radiological signs, medical conditions), which similarly demonstrated high rates of error propagation.^4,8,17-20^ Our results are also consistent with other studies of LLM benchmarks for medication-related tasks.^21-26^ In those studies, LLMs struggled with tasks such as matching drug names with available doses or identifying drug-drug interactions, tasks that are expected to be completed with high proficiency in the first year of the Doctor of Pharmacy curriculum.^21-26^ Ultimately, this type of evaluation may serve as a valuable early benchmark or checkpoint in the pre-clinical development of AI systems to help ensure safer deployment in patient care settings.

Mitigation prompts reduced confabulation rates considerably, indicating that the problem is a behavioral tendency to generate rather than a true knowledge deficit. Indication confabulations were nearly eliminated by mitigation, but dosing confabulations persisted, suggesting that dosage generation elicits deeper failure modes that are less amenable to simple instruction-based defenses. Setting model temperature to 0.0 (i.e., deterministic decoding) did not meaningfully reduce confabulation rates for most LLMs, and similar error patterns were observed across runs. These findings suggest that confabulations reflected systematic model behaviors rather than artifacts of sampling stochasticity. The use of a RAG approach further reduced confabulation rates and resulted in more uniform performance across models. Even so, residual confabulations persisted across all interventions, highlighting the need for multi-layered and complementary approaches to improving LLM reliability in clinical applications. While it appears that there may be solutions to solving this safety issue, it is important to emphasize that a responsible clinician, when faced with an unrecognized medication, would not make assumptions without subsequent information gathering (e.g., a Web search to learn about a newly available medication or clarifying with the patient or their pharmacy about the medication list).^27^ In clinical practice, recognizing uncertainty and using it as a trigger to engage in critical thinking, help-seeking, and evidence-gathering behaviors are essential for making safe treatment decisions.^6,22,28^ This concept is reflected in where human learners are taught well-defined core competencies and assessed against observable entrustable professional activities.^29^ Applying such a development and evaluation framework to AI systems can ensure their safety and efficacy in the clinical workplace,^30-33^ akin to licensing of medical practitioners.^34^

The majority of errors observed were inherited confabulations, in which models failed to identify the presence of a fabricated medication in the medication list and generated plausible but incorrect outputs. Epistemic confabulations, a pattern completion behavior in which a real medication used in clinical practice was substituted for the unrecognized fabricated medication, occurred less frequently but still produced clinically dangerous information. For example, the Pokémon “Loudred” was substituted with “lorazepam 1000 mg IV once,” while “Minior” was replaced by “minoxidil PO 110mg daily.” Both incidents represent potentially catastrophic dosing errors. This behavior indicates limited capacity for recognizing uncertainty or invalid inputs in medication data. In short, the LLMs do not say “I don’t know.”^6^ The implications of this finding in clinical practice are significant, as misspelled or phonetically rendered medication names could produce significant medication errors. Real-world data used for training will inevitably include errors: one study found that 21% of patients who read their personal electronic health record identified errors, of which 42% were deemed serious.^35^ Encountering poisoned data at the time of model training, whether through malicious attacks or unintentional yet routine errors that occur in healthcare delivery and communication, may lead to incorporation of that poisoned data into the LLM reasoning process.^17,36^ Beyond this, regulatory bodies such as the FDA and the Institute of Safe Medication Practices routinely emphasize the risks posed by look-alike and sound-alike medication names and highlight the need for safeguards to reduce inadvertent substitutions.^37^ Yet, LLMs were designed to generate answers and were specifically optimized through reinforcement learning from human feedback (RLHF) to improve alignment with human preferences.^38,39^ While RLHF has many strengths, it may also promote sycophancy (i.e., excessive agreeableness despite limited factual basis) and overly confident responses even in the setting of uncertainty.^38-40^ Combined with the propensity to confabulate, these behaviors raise substantial safety concerns for the use of LLMs in healthcare, and task-specific performance highlights the need to evaluate AI models using a battery of benchmarking tests mapped to observable tasks..^8,17,21,41^ The development of benchmarks that can stand as a bulwark against these tendencies may prove to be an essential domain for health AI.

This study has several limitations. We did not evaluate clinician performance in identifying fabricated medications and therefore cannot directly compare model and human responses; however, the ability to recognize when a medication is unfamiliar or potentially invalid is a core competency in health degree programs. The use of Pokémon characters as fabricated medication names represents a controlled but synthetic test condition that may not fully capture the complexity of real-world clinical inputs, where invalid inputs may be more subtle or context dependent. Each medication list contained four to six medications and exactly one fabricated medication, which underestimates the complexity of medication regimens and potential errors encountered in clinical practice, and future evaluations may benefit from more complex cases.^42-45^ All medication lists contained a fabricated medication, preventing evaluation of false positives. Model responses were constrained to a structured output format which, while ensuring consistency of outputs, did not allow for models to state “I don’t know” or ask followup questions. Evaluated strategies represent only a subset of potential approaches intended to improve LLM performance. Outputs were evaluated using an LLM-as-judge approach, which may introduce bias despite clinician validation of a random sample of outputs. Finally, although multiple leading contemporary LLMs were evaluated, there is a possibility other models may have had higher performance as reasoning models were not tested in this evaluation. Ultimately, additional strategies to reduce the consistently high confabulation rates are required, with this study serving as a proposed benchmark to test such future development.

This study provides two important additions to the understanding of LLMs for medication tasks. First, LLMs pose safety concerns without cautious use by experts. Second, the developed datasets and prompts may serve as a test to benchmark future LLMs and related training strategies to assess safe performance.

## Conclusion

In an evaluation of contemporary LLMs, confabulation rates in the presence of fabricated medications were high. Prompt-based mitigation strategies substantially reduced errors, but clinically concerning confabulations persisted across most models and tasks. These findings indicate that current LLMs may not meet consistently meet foundational safety expectations in medication-related applications. Further model development and standardized evaluation frameworks that explicitly test robustness to invalid inputs are essential for safe use in medication-related tasks.

## Data Availability

All data is publicly available in github at https://github.com/AIChemist-Lab/Pokemon-Drugs-Names.

https://github.com/AIChemist-Lab/Pokemon-Drugs-Name

